# Persistent circulating SARS-CoV-2 spike is associated with post-acute COVID-19 sequelae

**DOI:** 10.1101/2022.06.14.22276401

**Authors:** Zoe Swank, Yasmeen Senussi, Galit Alter, David R. Walt

**Affiliations:** Harvard Medical School, Boston, MA, USA; Brigham and Women’s Hospital, Department of Pathology, Boston, MA, USA; Wyss Institute for Biologically Inspired Engineering, Harvard University, Boston, MA, USA; Ragon Institute of MGH, MIT and Harvard, Camrbridge, MA, 02139; Division of Infectious Diseases, Massachusetts General Hospital, Boston, 02114

## Abstract

The diagnosis and management of post-acute sequelae of COVID-19 (PASC) poses an ongoing medical challenge. Identifying biomarkers associated with PASC would immensely improve the classification of PASC patients and provide the means to evaluate treatment strategies. We analyzed plasma samples collected from a cohort of PASC and COVID-19 patients (n = 63) to quantify circulating viral antigens and inflammatory markers. Strikingly, we detect SARS-CoV-2 spike antigen in a majority of PASC patients up to 12 months post-diagnosis, suggesting the presence of an active persistent SARS-CoV-2 viral reservoir. Furthermore, temporal antigen profiles for many patients show the presence of spike at multiple time points over several months, highlighting the potential utility of the SARS-CoV-2 full spike protein as a biomarker for PASC.

## Introduction

While symptoms resulting from infection with severe acute respiratory syndrome coronavirus 2 (SARS-CoV-2) typically resolve within a few weeks, some individuals experience persistent symptoms following the acute phase of coronavirus disease (COVID-19). The associated syndrome, termed post-acute sequelae of COVID-19 (PASC) or long COVID, encompasses a range of symptoms, including, but not limited to, fatigue, anosmia, memory loss, gastrointestinal distress, and shortness of breath.^1−7^ Estimates vary as to the prevalence of PASC,^8−11^ but the World Health Organization (WHO) reports around one quarter of individuals with COVID-19 continue to experience symptoms 4-5 weeks after a positive test and approximately one in 10 have continuing symptoms after 12 weeks.^12^

Although recent studies provide some clues, the underlying etiology of PASC remain hypotheses.^13^ This lack of mechanistic clarity is due partly to the heterogeneity of patient recruitment and inconsistencies in defining PASC patients, making it difficult to integrate results across studies. Disentangling the complex biology of PASC will rely on the identification of biomarkers that enable classification of patient phenotypes. Here, we analyze plasma samples collected from PASC and COVID-19 patients to determine the levels of SARS-CoV-2 antigens and cytokines and identify a blood biomarker that appears in the majority of PASC patients.

## Results

We analyzed plasma samples from a cohort of 63 individuals previously infected with SARS-CoV-2, 37 of whom were diagnosed with PASC. For most of the PASC patients (n= 31), blood samples were collected two or more times up to 12 months after their first positive result with either a nasopharyngeal swab RT-PCR test or an anti-SARS-CoV-2 antibody test. Blood was also collected from individuals who suffered from COVID-19, but were not diagnosed with PASC, up to five months post-diagnosis. The majority of PASC patients were female (n = 30), consistent with other studies that found women are predominantly affected by persistent symptoms following SARS-CoV-2 infection.^11,14−16^ Of the individuals who were not diagnosed with PASC (n = 26), ten were admitted to the intensive care unit (ICU) and seven were intubated. A complete overview of patient characteristics is provided in Supplementary Table 1.

Using previously developed and optimized ultra-sensitive single molecule array (Simoa) assays, we measured the concentration of SARS-CoV-2 antigens, including the S1 subunit of spike, full length spike, and nucleocapsid (N), in the collected plasma samples (Figure 1).^17−19^ We were able to detect either S1, spike or N in approximately 65% of the patients diagnosed with PASC at any given time point, several months after SARS-CoV-2 infection. Out of all three antigens measured, we detected spike most often in 60% of the PASC patients, whereas we did not detect spike at all in the COVID-19 patients. S1 was detected to a lesser degree in about one-fifth of the PASC patients and N was detected in a single patient at multiple time points. In line with our previous work,^18^ we detected S1 and N in COVID-19 patients, often those hospitalized with severe disease and within the first week post-diagnosis but we did not detect the full spike protein in any of these patients.

**Figure 1.**
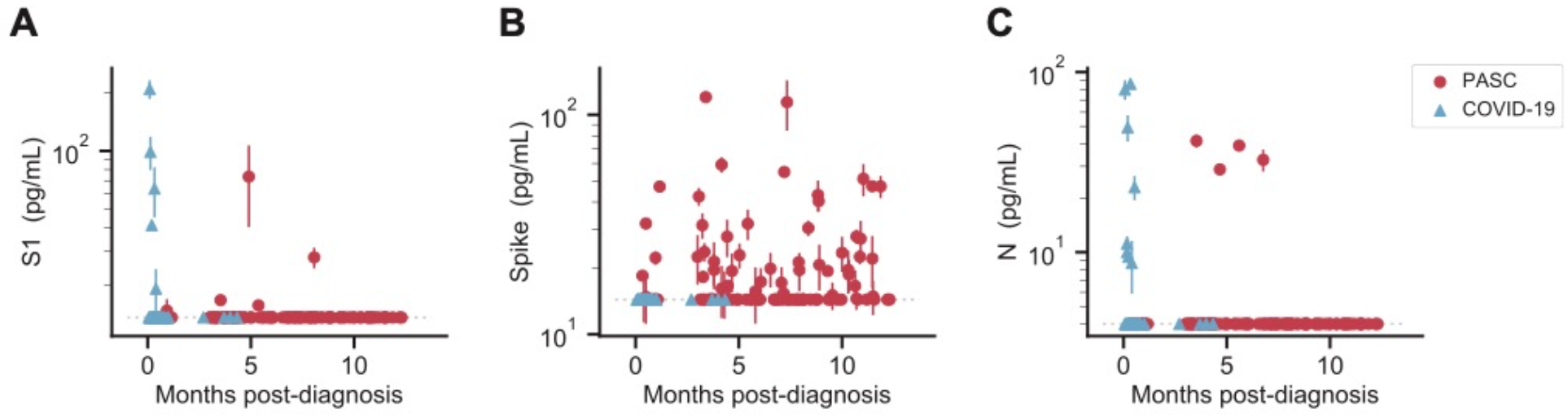
SARS-CoV-2 antigen levels versus time. The concentration of S1 **(A)**, spike **(B)**, and N **(C)** measured in the plasma of individuals over time after diagnosis with PASC or COVID-19 following SARS-CoV-2 infection. Multiple data points may correspond to the same individual, where repeat sampling was available. Data points represent mean values ± SD (n = 2). Dashed lines indicate the LOD for each assay.

A number of PASC patients and a subset of COVID-19 patients blood samples were collected at multiple time points, allowing us to analyze the change in antigen levels over time. Of the PASC patients for which we had longitudinal samples, we detected antigen more than once in 12 patients, reinforcing the persistent presence of circulating antigen (Figure 2). In only a few cases, the presence of S1 or spike may be correlated with vaccination, however according to our previous findings, S1 is only detected within the first two weeks after the first dose and spike is rarely detected.^20^ Most significantly, we observe patterns of sustained full spike antigen levels over the course of several months in many patients. In other cases, we observe fluctuations between antigen being detected and not detected, indicating that the time of sampling is important. In contrast, temporal antigen profiles of six COVID-19 patients show high antigen levels soon after diagnosis, quickly dropping below the limit of detection (Figure S1).

**Figure 2.**
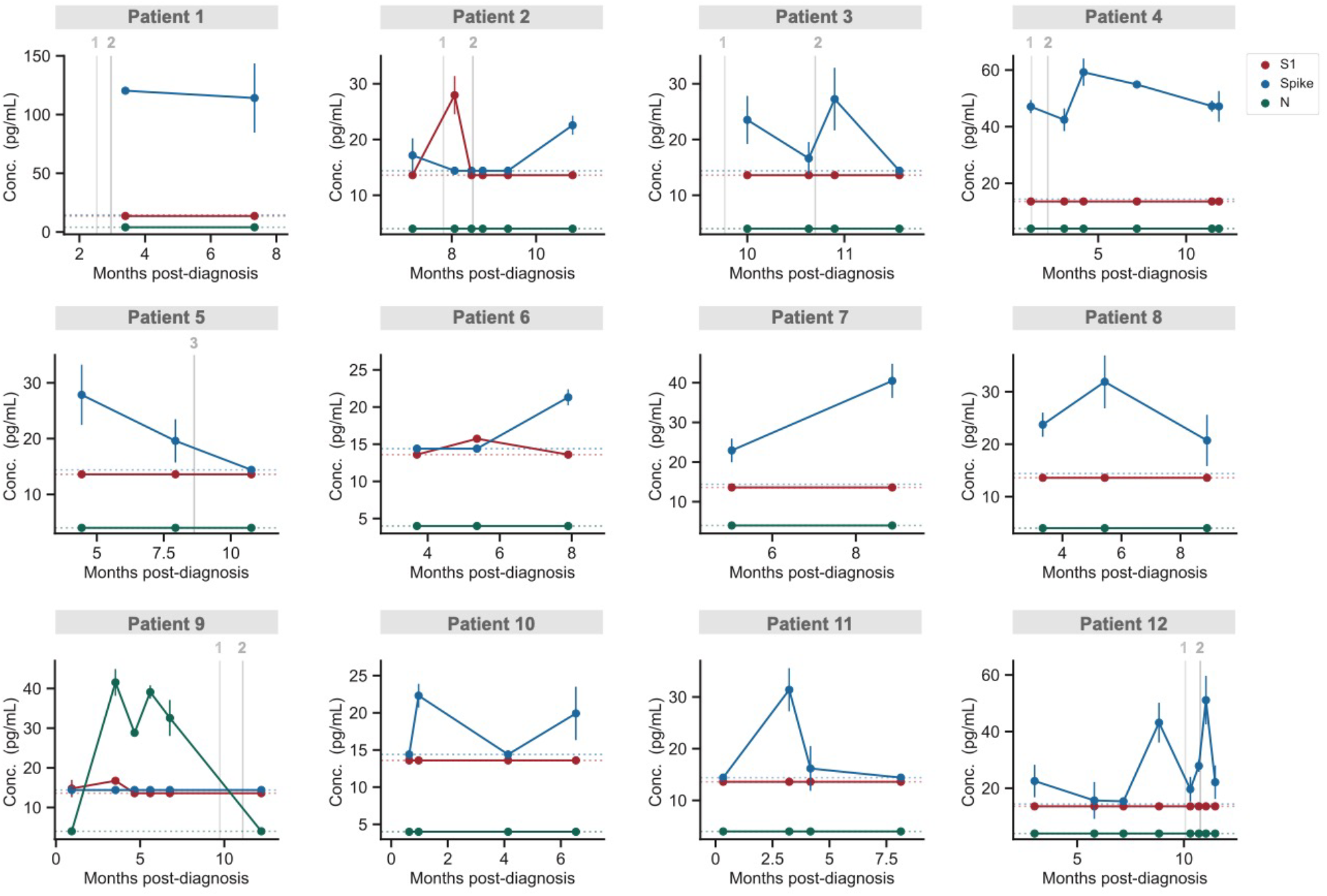
Temporal profiling of SARS-CoV-2 antigens for individual PASC patients. S1, spike, and N levels versus time for 12 patients diagnosed with PASC. Data points represent mean values ± SD (n = 2). Grey vertical lines correspond to the times when each patient received either the first, second or third dose of a COVID-19 vaccine. If an individual was not vaccinated within one month of the time frame shown, no vaccination information is shown. Dashed horizontal lines indicate the LOD for each assay.

We also measured the plasma concentrations of a panel of cytokines, including IFN-γ, IL-1β, IL-4, IL-5, IL-6, IL-8, IL-10, IL-12p70, IL-22, and TNF-⍺, but there were no clear findings (Figure S2, S3).

## Discussion

With the goal of discovering biomarkers that correlate with PASC, we measured the concentration of SARS-CoV-2 antigens and cytokines in plasma samples collected from a cohort of PASC and COVID-19 patients. Most notably, we detected circulating SARS-CoV-2 spike in the majority of PASC patients, but in none of the COVID-19 patients. Although our sample size is small, the detection of spike at multiple time points 2-12 months after infection is compelling. Temporal fluctuations in antigen levels for a subset of individuals indicates the importance of longitudinal sampling as we may not have detected antigen in some cases if blood was collected at a single time point. Furthermore, those who volunteer to participate in PASC studies tend to have been hospitalized with severe COVID-19, notwithstanding that the majority of those who acquire SARS-CoV-2 and develop PASC are not hospitalized.^13^ The minority of our PASC cohort was hospitalized (n=2), suggesting that our findings relate to SARS-CoV-2 infection rather than effects that arise after severe illness and hospitalization.

The presence of circulating spike supports the hypothesis that a reservoir of active virus persists in the body. Another preliminary study found SARS-CoV-2 RNA in multiple anatomic sites up to seven months after symptom onset, corroborating viral antigen persistence. Post-mortem tissue analysis revealed the presence of SARS-CoV-2 RNA and viral specific protein expression not only in respiratory tissues, but also in cardiovascular, lymphoid, gastrointestinal, peripheral nervous, and brain tissues in the majority of individuals analyzed (n = 44).^21^ Furthermore, in our previous study, a reservoir of replicating SARS-CoV-2 was found to occur in the gastrointestinal tract of children who develop multisystem inflammatory syndrome (MIS-C). In that study, we detected elevated levels of circulating spike weeks after initial SARS-CoV-2 infection, due to leakage from the gut into the blood stream.^22^ In another study, analysis of the gut microbiome up to six months after SARS-CoV-2 infection revealed alterations in the gut microbiome composition that correlated with PASC,^23^ suggesting that certain conditions or predispositions may foster the persistence of replicating viral reservoirs.

If viral reservoirs persist in the body of PASC patients, then why do we not also detect circulating N in a larger proportion of patients? It is possible that N is preferentially hydrolyzed, whereas spike may be more efficiently transported into the bloodstream, evading degradation. Alternatively, circulating anti-N antibodies may be more effective at clearing N compared to the anti-spike antibodies produced. Furthermore, PASC is a heterogeneous syndrome, possibly dependent on the tissue in which the viral reservoir persists. This heterogeneity may have an effect on whether spike or N is then detected; future investigations are necessary to resolve these unanswered questions.

Persistent viral reservoirs are a likely cause of PASC symptoms, but circulating spike may also give rise to symptoms. Similar to bacterial superantigens, SARS-CoV-2 spike protein contains structural motifs that strongly interact with T cell receptors, creating an imbalance in the T cell receptor repertoire.^24,25^ Unique to SARS-CoV-2, it is thought that the superantigen-like motif may account for the hyperinflammatory response observed in severe COVID-19 and MIS-C patients. Although spike does not instigate a cytokine storm in PASC patients, it could act through other mechanisms that interfere with normal cellular function. For instance, spike alone was shown to disrupt signaling pathways and induce dysfunction in pericytes, vascular endothelial cells, and the blood brain barrier.^26−29^ Alternatively, PASC may manifest as cellular exhaustion or the induction of refractory inflammatory response − but with persistent tissue localized dysfunction.

If PASC is the result of persistent viral replication in tissues outside the lung, it may also explain why changes in circulating inflammatory markers may be subtle when measured in blood, although they may be elevated within a given tissue. Other work indicates a trend towards elevated IL-6 and TNF-⍺ in PASC patients, although fold-changes compared to controls are small.^30,31^ In a different study, the combination of inflammatory mediators, including IL-6, IFN-γ, IFN-β, PTX3, and IFN-λ2/3, was found to be a more informative predictor of PASC.^32^ Therefore, even though we do not observe cytokine levels to be elevated above the normal range in most cases, monitoring cytokines over time in a larger cohort warrants further investigation. Furthermore, the absence of any distinguishing serological features in our PASC cohort may be due to the late sampling times post infection. Lower titers of IgM and IgG3 measured during acute infection correlate with the development of PASC,^33^ but over time many immunological signatures may diminish,^34^ emphasizing the importance of analyzing patient samples near the time of primary infection and over the long-term.

In conclusion, the presence of circulating spike in PASC patients up to 12 months post-diagnosis strongly suggests that SARS-CoV-2 viral reservoirs persist in the body. Additionally, the detection of spike in a majority of individuals included in our PASC cohort provides strong support for the use of spike as a biomarker for PASC. If PASC patients can be identified based on the direct measurement of spike protein, characterizing patient phenotypes and assessing treatment strategies will become increasingly effective.

## Data Availability

All data produced in the present study are available upon request to the authors.

## Acknowledgments

Funding for this work came from a generous donation from Barbara and Amos Hostetter. We thank the MassCPR PASC consortium and the Ragon Institute/MassCPR Biobank team for financial and sample processing support.

## Materials and Methods

### Sample Collection

All subjects had a PCR confirmed COVID-19 infection and were included in our large Massachusetts Coalition for Pathogen Readiness (MassCPR) consortium biorepository program at the Ragon Institute. Samples for this study were selected based on reported symptoms captured in routine visit questionnaires. Individuals that reported symptoms for over 30 days after COVID-19 were included in our PASC cohort and included individuals reporting 1-5 protracted symptoms including anosmia, fatigue, brain-fog, persistent coughing, persistent headaches, etc. Samples were collected serially up to 360 days following PCR confirmed SARS-CoV-2 infection. In addition, non-PASC controls included PCR confirmed COVID-19 convalescents with no reported prolonged symptoms sampled after diagnosis. All individuals gave informed consent and the study was approved by the Massachusetts General Brigham Hospital. Plasma samples from 16 additional COVID-19 patients were obtained from the Mass General Brigham Biobank. Patient demographics and clinical information were extracted from electronic medical records. Analysis of COVID-19 plasma samples was approved by the Institutional Review Board of Brigham and Women’s Hospital.

### Simoa SARS-CoV-2 antigen assays

SARS-CoV-2 S1, spike, and N antigens were measured in plasma samples using previously developed single molecule array (Simoa).^18,19^ Plasma samples were first incubated with dithiothreitol (DTT) and protease inhibitors (Halt Protease Inhibitor Cocktail, ThermoFisher) at 37 °C for 15 minutes to denature any bound antibodies. The samples were then diluted 8-fold in a sample diluent buffer (Quanterix Corp.) and assays were run using an HD-X Analyzer (Quanterix Corp.).

### Cytokines

Cytokine levels were measured in plasmas samples using the CorPlex Cytokine Panel (Quanterix Corp). Plasma samples were diluted 4-fold in sample diluent buffer and assays were performed following the CorPlex manufacturer protocols. Each CorPlex cytokine panel kit was analyzed by the SP-X Imaging and Analysis System (Quanterix Corp.).

---

**Supplementary Table 1.** Patient characteristics for the PASC and COVID-19 cohorts.

|  | PASC (n = 37) | COVID-19 (n = 26) |
| --- | --- | --- |
| Age, median (range) | 46 (27, 70) | 63 (26, 83) |
| Sex, number (%) |  |  |
| Female | 30 (81%) | 10 (38%) |
| Male | 7 (19%) | 16 (62%) |
| Race or ethnic group, number |  |  |
| Asian | 0 | 1 |
| Black or African American | 1 | 5 |
| Hispanic | 1 | 2 |
| White | 35 | 18 |
| ICU admission, number | 2 | 10 |
| Intubation, number | 2 | 7 |

**Figure S1.**
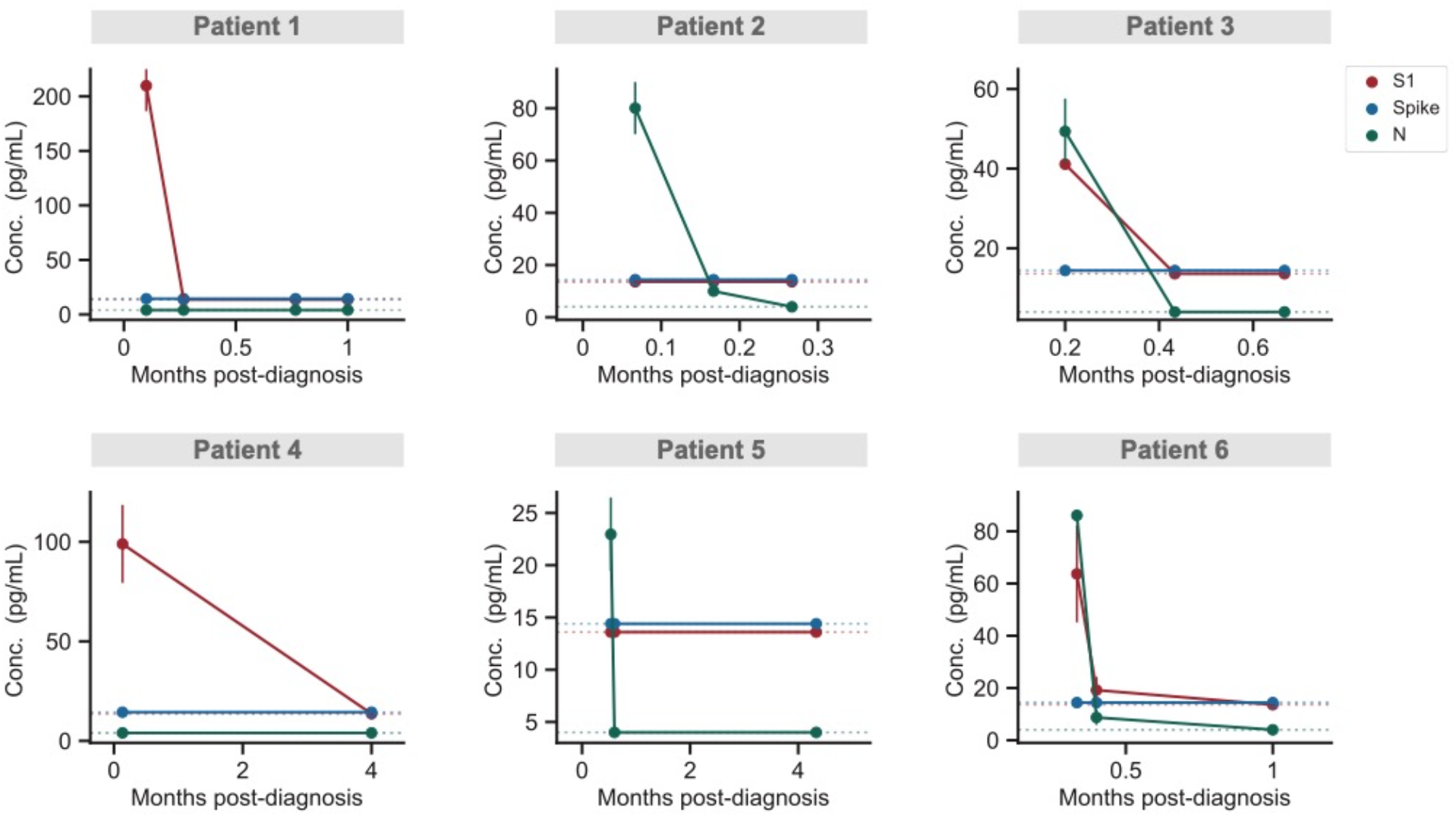
Temporal profiling of SARS-CoV-2 antigens for individual COVID-19 patients. S1, spike, and N levels versus time for six COVID-19 patients diagnosed with PASC. Data points represent mean values ± SD (n = 2). Dashed horizontal lines indicate the LOD for each assay.

**Figure S2.**
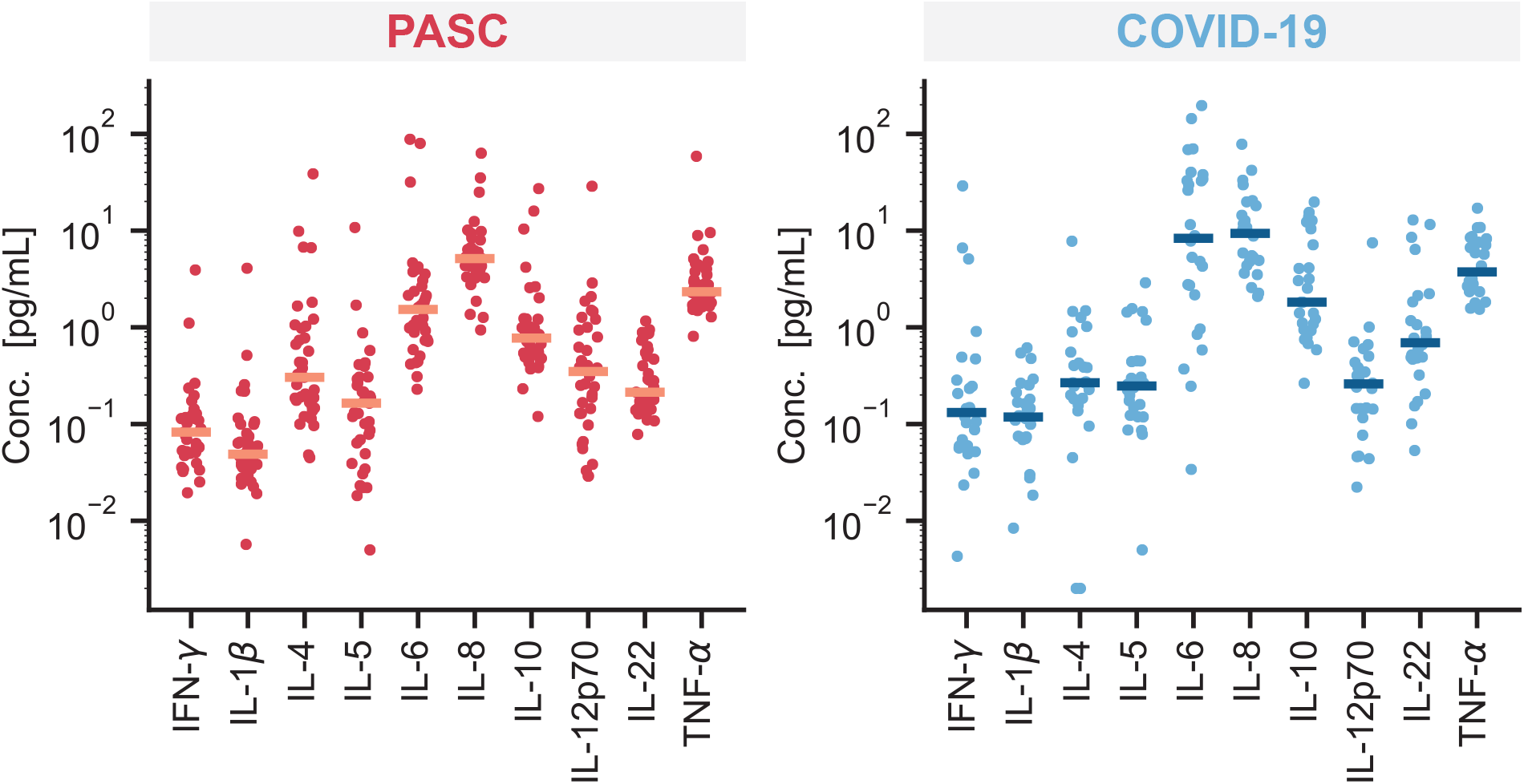
Cytokine levels in PASC and COVID-19 patients. Measured concentration of a panel of cytokines in plasma samples collected from PASC and COVID-19 patients. If longitudinal sampling was performed, the maximal value is plotted. Data points represent mean values (n = 2) and horizontal bars indicate the median concentrations for each cytokine.

**Figure S3.**
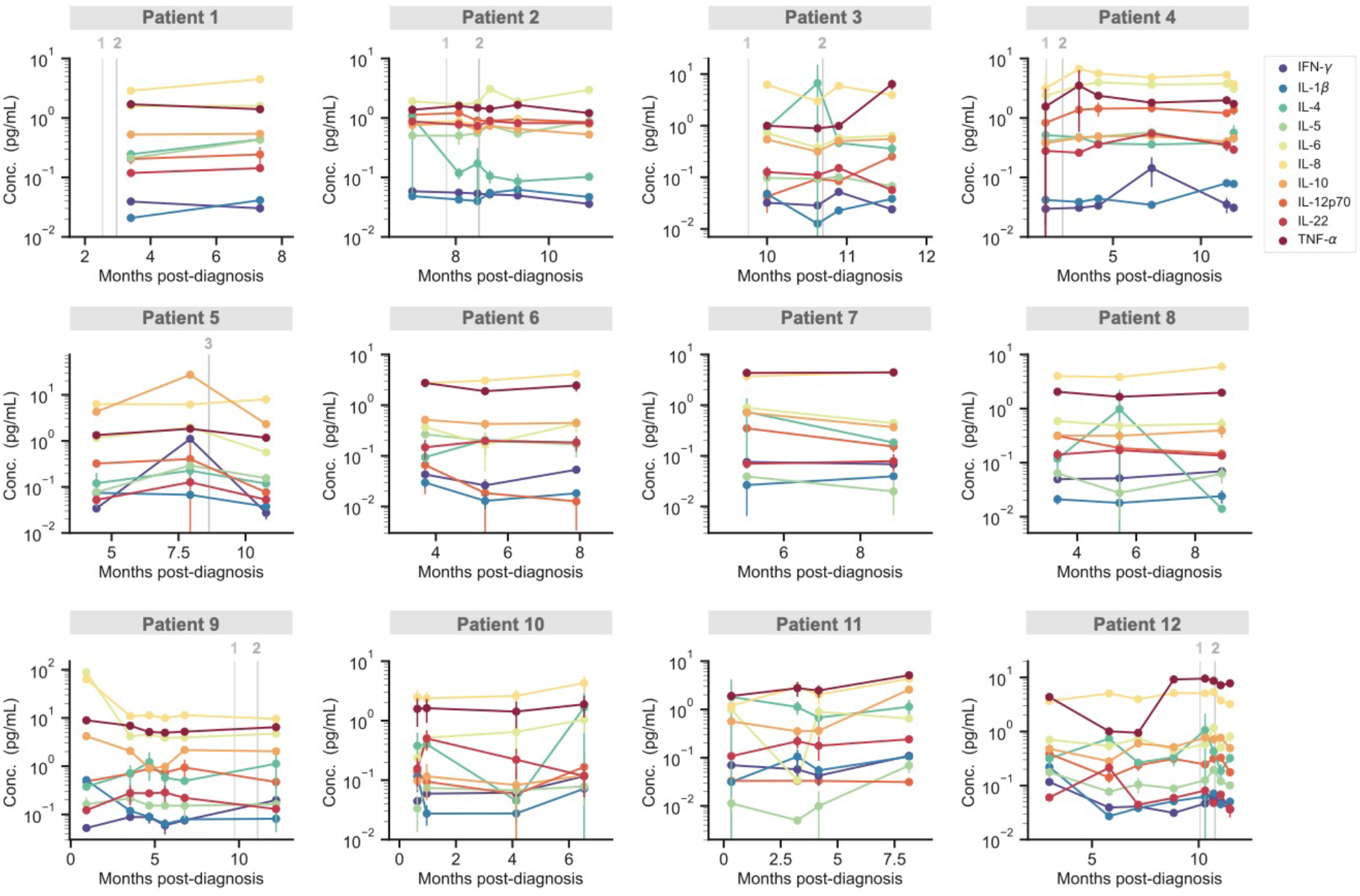
Temporal profiling of cytokines for individual PASC patients. The concentration of 10 cytokines measured over time for 12 individual PASC patients, corresponding to the same patients shown in Figure 2. Grey vertical lines correspond to the times when each patient received either the first, second or third dose of a COVID-19 vaccine. Data points represent mean values ± SD (n = 2).

